# Analysis across Taiwan Biobank, Biobank Japan and UK Biobank identifies hundreds of novel loci for 36 quantitative traits

**DOI:** 10.1101/2021.04.12.21255236

**Authors:** Chia-Yen Chen, Tzu-Ting Chen, Yen-Chen Anne Feng, Ryan J. Longchamps, Shu-Chin Lin, Shi-Heng Wang, Yi-Hsiang Hsu, Hwai-I Yang, Po-Hsiu Kuo, Mark J. Daly, Wei J. Chen, Hailiang Huang, Tian Ge, Yen-Feng Lin

**Affiliations:** Biogen, Cambridge, MA, USA; Psychiatric and Neurodevelopmental Genetics Unit, Massachusetts General Hospital, Boston, MA, USA; Analytic and Translational Genetics Unit, Massachusetts General Hospital, Boston, MA, USA; Stanley Center for Psychiatric Research, Broad Institute of MIT and Harvard, Cambridge, MA, USA; Center for Neuropsychiatric Research, National Health Research Institutes, Miaoli, Taiwan; Vertex Pharmaceuticals, Boston, MA, USA; Department of Occupational Safety and Health and Department of Public Health, College of Public Health, China Medical University, Taichung, Taiwan; Marcus Institute for Aging Research and Harvard Medical School, Boston, MA, USA; Beth Israel Deaconess Medical Center, Boston, MA, USA; Harvard School of Public Health, Boston, MA, USA; Broad Institute of MIT and Harvard, Cambridge, MA, USA; Genomics Research Center, Academia Sinica, Taipei, Taiwan; Institute of Clinical Medicine, National Yang-Ming University, Taipei, Taiwan; Graduate Institute of Medicine, College of Medicine, Kaohsiung Medical University, Kaohsiung, Taiwan; Biomedical Translation Research Center, Academia Sinica, Taipei, Taiwan; Department of Public Health & Institute of Epidemiology and Preventive Medicine, College of Public Health, National Taiwan University, Taipei, Taiwan; Department of Psychiatry, College of Medicine and National Taiwan University Hospital, Taipei, Taiwan; Institute for Molecular Medicine Finland FIMM, University of Helsinki, Helsinki, Finland; Department of Medicine, Harvard Medical School, Boston, MA, USA; Department of Psychiatry, Massachusetts General Hospital, Harvard Medical School, Boston, MA, USA; Department of Public Health & Medical Humanities, School of Medicine, National Yang Ming Chiao Tung University, Taipei, Taiwan; Institute of Behavioral Medicine, College of Medicine, National Cheng Kung University, Tainan, Taiwan

**Author notes:** Correspondence to: Yen-Feng Lin; Tian Ge; Hailiang Huang; Yen-Chen Anne Feng; Chia-Yen Chen. These authors contributed equally (Chia-Yen Chen, Tzu-Ting Chen, Yen-Chen Anne Feng, Ryan J. Longchamps, Shu-Chin Lin).

## Abstract

Genome-wide association studies (GWAS) have identified tens of thousands of genetic loci associated with human complex traits and diseases^1,2^. However, the majority of GWAS were conducted in individuals of European ancestry^3^. Failure to capture global genetic diversity has limited biological discovery and impeded equitable delivery of genomic knowledge to diverse populations^4^. Here we report findings from 102,900 individuals across 36 human quantitative traits in the Taiwan Biobank (TWB), a major biobank effort that broadens the population diversity of genetic studies in East Asia (EAS). We identified 979 novel genetic loci, pinpointed novel causal variants through fine-mapping, compared the genetic architecture across TWB, Biobank Japan (BBJ)^5–7^ and UK Biobank (UKBB)^8,9^, and evaluated the utility of cross-phenotype, cross-population polygenic risk scores (PRS) in disease risk prediction. These results demonstrated the potential to advance genomic discovery through increasing and diversifying GWAS populations, and provided insights into the common genetic background for human complex traits in East Asian populations.

## Main

The Taiwan Biobank (TWB) is a community-based prospective cohort study of the Taiwanese population with multi-omics genomic data, and longitudinal phenotypic and environmental measures (see https://www.twbiobank.org.tw/new_web_en/ for more information). Genotyping was performed using two different customized genome-wide arrays^10^. A total of 27,719 participants genotyped on the TWBv1 array and 83,207 participants genotyped on the TWBv2 array were included in this study and subsequently went through genotype quality control (QC) and imputation. Figure 1 provides an overview of the TWB samples, the traits examined, their abbreviations, and the analyses conducted in this study.

**Figure 1:**
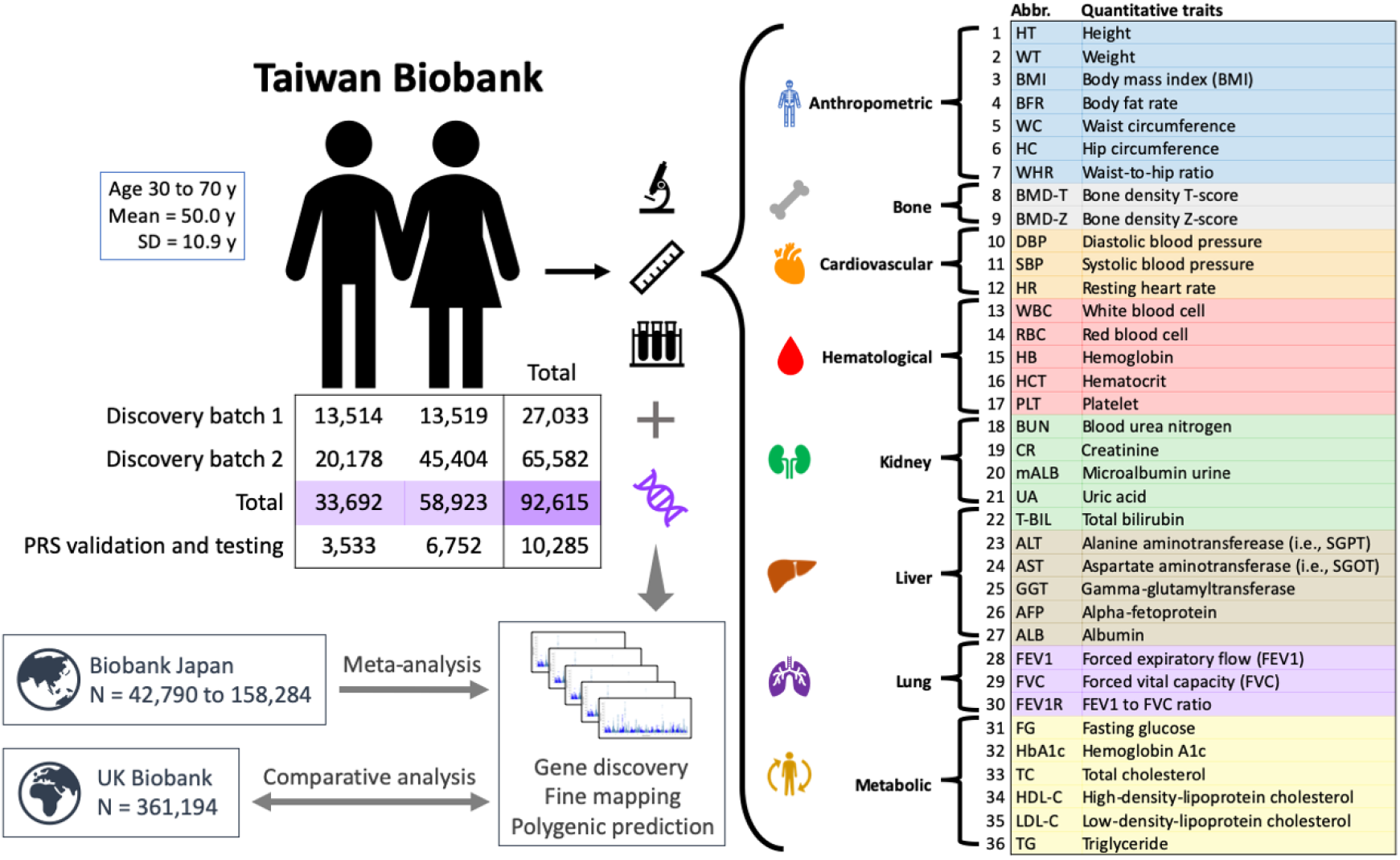
Overview of the Taiwan Biobank sample and analysis. The abbreviations and index numbers for the 36 quantitative traits examined in this study are used throughout the text, tables and figures. The sample size noted in the figure reflects the final analytical sample size after genotype quality control and imputation.

After stringent QC, we performed GWAS on 36 quantitative traits (Figure 2a; Supplementary Tables 1 & 2) in 92,615 individuals with imputed genotype data across the two genotyping arrays (Methods). We selected these traits as they are intermediate phenotypes relevant to an individual’s health and disease status. We used Regenie^11^, a two-step whole genome regression method for genetic association tests that accounts for sample relatedness and population structure, to perform association analyses on the two discovery batches (27,033 and 65,582 individuals) separately. LD score regression (LDSC)^12^ intercept, λ_GC_ and λ_1000_ showed that there was negligible inflation due to population stratification in these GWAS (Supplementary Table 3). All traits had highly consistent genetic architectures across the two discovery batches, as shown by the high between-batch genetic correlations (r_g_) estimated by LDSC^13^ (median = 1.028; Supplementary Table 3). We then meta-analyzed discovery batch 1 and 2 GWAS using an inverse-variance-weighted fixed-effect approach^14^. Using FUMA^15^ with the 1000 Genomes Project (1KG)^16^ phase 3 EAS samples as the LD reference, we identified 1,907 independent genome-wide significant loci (*P*-value < 5×10^−8^) across the 36 traits (Supplementary Tables 1 & 2), among which 1,287 loci survived Bonferroni correction for the number of traits tested (*P*-value < 5×10^−8^/36). The number of genome-wide significant loci per trait ranged from 1 for FEV1 and FEV1 to FVC ratio (FEV1R), to 211 for height (HT). Using LDSC^12,13^, we estimated the SNP-based heritability (h^2^_g_) for each trait (Figure 2b; Supplementary Table 4), which ranged from 0.009 (FEV1R) to 0.384 (HT), and pairwise r_g_ between these traits (Figure 2c; Supplementary Table 5), which identified clusters of highly genetically correlated traits (e.g., BFR, BMI, WT, WC, HC and WHR).

**Figure 2:**
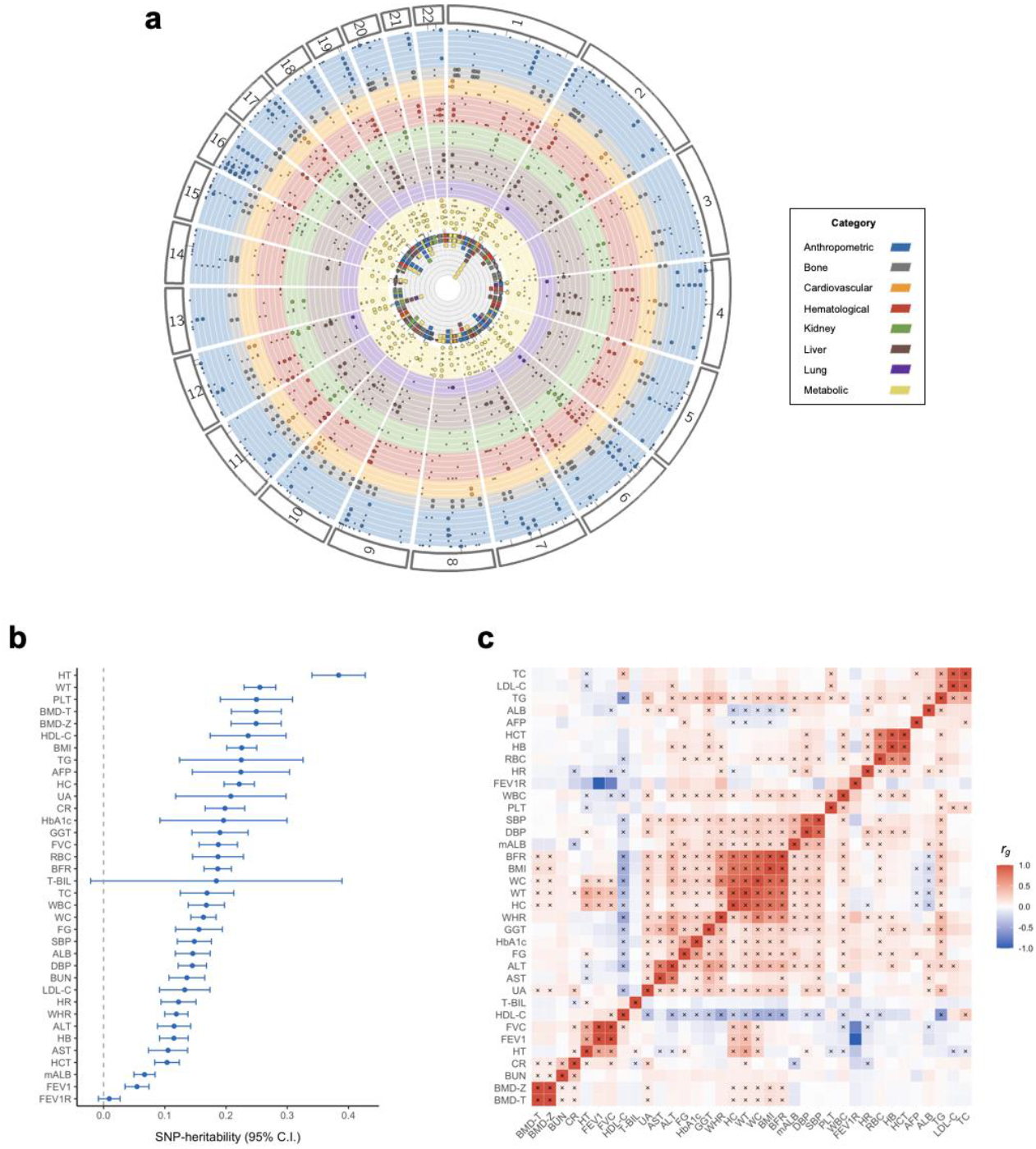
GWAS results for 36 quantitative traits in the Taiwan Biobank. **a** A Fuji plot summary of genome-wide significant loci associated with the 36 traits identified by Regenie. Each layer of the plot represents a single trait, with traits within the same category grouped by the same color. Each dot on the circular layouts represents a genome-wide significant hit; trait-specific associations are shown in a smaller-sized dot, while cross-trait associations are shown in a larger-sized dot. The inner circle summarizes all significant associations across traits into a stacked, circular plot. **b** SNP-based heritability (h^2^_g_) for the 36 traits in TWB estimated using univariate LD score regression (LDSC). Abbreviations of the traits are listed in Figure 1. The complete set of h^2^_g_ estimates, including standard errors and p-values, is available in Supplementary Table 4. The unusually large confidence interval (CI) of the h^2^_g_ estimate for total bilirubin (T-BIL) is driven by a Mendelian locus on chromosome 2, harboring the *UGT1A1* gene. Modeling the signal in this locus as a fixed effect and removing the locus from the LDSC analysis produced a similar point estimate of h^2^_g_ with a much smaller CI (see Methods). **c** Pairwise genetic correlations (r_g_) between the 36 traits in TWB estimated using bivariate LDSC. Significant r_g_ after FDR correction is indicated by a cross sign. The complete set of r_g_ estimates, including standard errors and p-values, is available in Supplementary Table 5. Both h^2^_g_ and r_g_ analyses were based on GWAS generated by linear regression (see Methods).

We fine-mapped genome-wide significant loci using SuSiE^17^ (Methods). Of the 1,907 loci, 1,615 were fine-mapped to a total of 1,972 credible sets, each representing an independent association signal (292 loci failed to identify a reliable credible set that passed QC thresholds; see Methods). Out of the 1,972 credible sets, 232 were mapped to a single variant with posterior inclusion probability (PIP) > 95%, among which 24 were missense variants (Table 1; Supplementary Table 6). This represented a 9.8-fold enrichment of missense variants (*P*-value < 1×10^−16^) compared with all variants in the fine-mapped loci, demonstrating the importance of missense variants in the quantitative traits examined in this study. All fine-mapped missense variants with the corresponding GWAS available in Biobank Japan (BBJ) and/or UK Biobank (UKBB) were replicated with the same direction of effect and study-wide significance (*P*-value < 0.05/15=0.0033). These putative causal missense variants not only replicated previous findings (e.g., T1412N in *CPS1* for PLT^18^ and V174A in *SLCO1B1* for T-BIL^19^), but also represented novel causal variants in well-known genes (e.g., R103W in *EXOC3L4* for GGT^20^) and implicated novel genes (e.g., D1171N in *RREB1* for BMD-T and BMD-Z). Some of these high-PIP missense variants were also highly pleiotropic (e.g., S267F in *SLC10A1* is associated with GGT, LDL-C and TC), suggesting their roles in multiple complex traits.

**Table 1:** Fine-mapped missense variants with posterior inclusion probability (PIP) > 0.95 in the Taiwan Biobank. Base pair position is in hg19. Allele freq (EAS) and Allele freq (EUR) are the allele frequencies of the reference allele (REF column) in East Asian and European populations, calculated from East Asians and non-Finnish Europeans in gnomAD v2.1.1, respectively. Marginal beta is the marginal effect size of the reference allele. BBJ and UKBB *P*-values for each variant were extracted from the BBJ and UKBB GWAS, respectively. No GWAS: GWAS of the trait has not been conducted in BBJ or UKBB. Missing: the fine-mapped missense variant and all variants in LD (*R*^*2*^ > 0.4) with the missense variant were not present in the GWAS. Low MAF: minor allele frequency < 0.1%. For BBJ, seven variants, including variants in LD (*R*^*2*^ > 0.4) with them, were not available in the GWAS summary statistics (labeled as “Missing”), due to limited genomic coverage of the 1000 Genomes Project phase 1 reference panel (unpublished data). For UKBB, seven variants had MAF < 0.1%, and thus were not included in the GWAS (labeled as “Low MAF”). Full fine-mapping results are available in Supplementary Table 6.

| Trait | Chr | Variant | Position | REF | ALT | Allele freq (EAS) | Allele freq (EUR) | Marginal beta | SE | P-value | PIP | BBJ P-value | UKBB P-value | Gene | Amino acid change |
| --- | --- | --- | --- | --- | --- | --- | --- | --- | --- | --- | --- | --- | --- | --- | --- |
| TG | 2 | rs1260326 | 27730940 | T | C | 0.492 | 0.591 | 0.137 | 0.005 | 7.80E-191 | 1.00 | 1.70E-94 | 2.50E-418 | *GCKR* | Leu256Pro |
| PLT | 2 | rs1047891 | 211540507 | A | C | 0.161 | 0.311 | -0.048 | 0.006 | 4.60E-14 | 0.98 | 1.80E-03 | 2.90E-44 | *CPS1* | Thr1412Asn |
| BMD-T | 6 | rs9379084 | 7231843 | A | G | 0.119 | 0.118 | -0.055 | 0.007 | 1.70E-14 | 0.99 | No GWAS | No GWAS | *RREB1* | Asp1171Asn |
| BMD-Z | 6 | rs9379084 | 7231843 | A | G | 0.119 | 0.118 | -0.055 | 0.007 | 1.10E-14 | 0.99 | No GWAS | No GWAS | *RREB1* | Asp1171Asn |
| FG | 6 | rs146340667 | 39040708 | A | G | 0.011 | 7.74E-06 | -0.145 | 0.022 | 8.90E-11 | 1.00 | Missing | Low MAF | *GLP1R* | Val194Ile |
| HbA1C | 7 | rs2233580 | 127253550 | T | C | 0.101 | 7.74E-06 | 0.098 | 0.008 | 2.60E-37 | 1.00 | Missing | Low MAF | *PAX4* | Arg192His |
| HT | 8 | rs12541381 | 135649848 | A | G | 0.300 | 0.250 | -0.039 | 0.005 | 1.30E-14 | 1.00 | 1.90E-11 | 3.40E-43 | *ZFAT* | Pro90Ser |
| LDL-C | 11 | rs11601507 | 5701074 | A | C | 0.090 | 0.077 | 0.059 | 0.008 | 3.90E-13 | 1.00 | 3.10E-04 | 4.40E-13 | *TRIM5* | Val112Phe |
| TC | 11 | rs11601507 | 5701074 | A | C | 0.090 | 0.077 | 0.054 | 0.008 | 3.30E-11 | 0.99 | 2.20E-03 | 2.70E-07 | *TRIM5* | Val112Phe |
| T-BIL | 12 | rs4149056 | 21331549 | C | T | 0.112 | 0.159 | 0.154 | 0.007 | 1.00E-96 | 1.00 | 6.20E-46 | 1.40E-638 | *SLCO1B1* | Val174Ala |
| HT | 12 | rs2277339 | 57146069 | G | T | 0.206 | 0.105 | -0.040 | 0.006 | 6.20E-12 | 1.00 | 1.80E-19 | 1.90E-10 | *PRIM1* | Asp5Ala |
| TC | 12 | rs1169288 | 121416650 | C | A | 0.337 | 0.357 | 0.044 | 0.005 | 2.80E-20 | 0.99 | 1.90E-07 | 2.00E-25 | *HNF1A* | Ile27Leu |
| GGT | 14 | rs2296651 | 70245193 | A | G | 0.095 | 1.55E-05 | 0.061 | 0.008 | 1.60E-14 | 0.97 | Missing | Low MAF | *SLC10A1* | Ser267Phe |
| LDL-C | 14 | rs2296651 | 70245193 | A | G | 0.095 | 1.55E-05 | -0.072 | 0.008 | 1.50E-19 | 1.00 | Missing | Low MAF | *SLC10A1* | Ser267Phe |
| TC | 14 | rs2296651 | 70245193 | A | G | 0.095 | 1.55E-05 | -0.072 | 0.008 | 1.00E-19 | 1.00 | Missing | Low MAF | *SLC10A1* | Ser267Phe |
| GGT | 14 | rs200884137 | 103576369 | T | C | 0.007 | 9.78E-06 | -0.322 | 0.028 | 9.80E-31 | 0.99 | Missing | Low MAF | *EXOC3L4* | Arg103Trp |
| HT | 15 | rs3817428 | 89415247 | G | C | 0.105 | 0.271 | -0.050 | 0.008 | 5.20E-11 | 1.00 | 1.30E-13 | 4.10E-56 | *ACAN* | Asp2335Glu |
| HDL-C | 16 | rs2303790 | 57017292 | G | A | 0.029 | 1.39E-04 | 0.490 | 0.014 | 1.70E-272 | 1.00 | 5.00E-198 | Low MAF | *CETP* | Asp394Gly |
| ALB | 17 | rs11552708 | 7462555 | A | G | 0.305 | 0.118 | 0.047 | 0.005 | 2.10E-20 | 1.00 | 7.90E-05 | 1.00E-17 | *TNFSF13* | Gly67Arg |
| TG | 19 | rs58542926 | 19379549 | T | C | 0.063 | 0.074 | -0.105 | 0.010 | 2.10E-28 | 1.00 | 9.10E-14 | 1.70E-127 | *TM6SF2* | Glu167Lys |
| HDL-C | 19 | rs429358 | 45411941 | C | T | 0.084 | 0.149 | -0.132 | 0.008 | 6.50E-56 | 1.00 | 2.40E-24 | 1.20E-127 | *APOE* | Cys156Arg |
| HDL-C | 19 | rs686335 | 54754989 | T | G | 0.480 | 0.176 | -0.029 | 0.005 | 3.80E-10 | 1.00 | Missing | 7.80E-14 | *LILRB5* | Glu549Ala |
| HDL-C | 20 | rs1800961 | 43042364 | T | C | 0.014 | 0.032 | -0.143 | 0.020 | 2.90E-13 | 1.00 | 1.70E-04 | 3.00E-95 | *HNF4A* | Thr139Ile |
| TC | 20 | rs1800961 | 43042364 | T | C | 0.014 | 0.032 | -0.115 | 0.020 | 5.60E-09 | 1.00 | 3.30E-06 | 2.20E-44 | *HNF4A* | Thr139Ile |

Prior studies have shown that complex traits and diseases are genetically correlated at different levels between EAS and European (EUR) populations^21–23^, but the genetic overlap within EAS populations has not been characterized yet. Leveraging existing GWAS summary statistics from BBJ and UKBB, we investigated the comparative genetic architecture of quantitative traits within EAS (TWB vs. BBJ), and between EAS and EUR populations (TWB vs. UKBB and BBJ vs. UKBB). Intriguingly, among the 21 traits for which GWAS were available across the three biobanks, h^2^_g_ estimates in TWB were comparable with those in UKBB, but consistently higher than the h^2^_g_ estimates in BBJ, except for height (Figure 3a; Supplementary Table 7), even though many r_g_ estimates between TWB and BBJ were indistinguishable from 1 (median = 0.933; Figure 3b & 3c). When comparing within-EAS and cross-population (EAS vs. EUR) genetic correlations, within-EAS TWB-BBJ r_g_ estimates were in general higher than TWB-UKBB r_g_ and BBJ-UKBB r_g_ estimates (Figure 3b & 3c; Supplementary Table 8). Despite these differences, we note that all within-and cross-population r_g_ estimates were high (median=0.927). Taken together, these results suggested that the genetic architecture for the quantitative traits examined here was largely consistent across EAS and EUR populations. The systematic differences in TWB vs. BBJ h^2^_g_ estimates and TWB-UKBB vs. BBJ-UKBB r_g_ estimates likely reflected differences in sample ascertainment. In particular, both TWB and UKBB are community- or population-based prospective cohorts, while BBJ is a multi-institutional hospital-based registry, in which biomarker measurements may be affected by the health condition and medication use of the patients.

**Figure 3:**
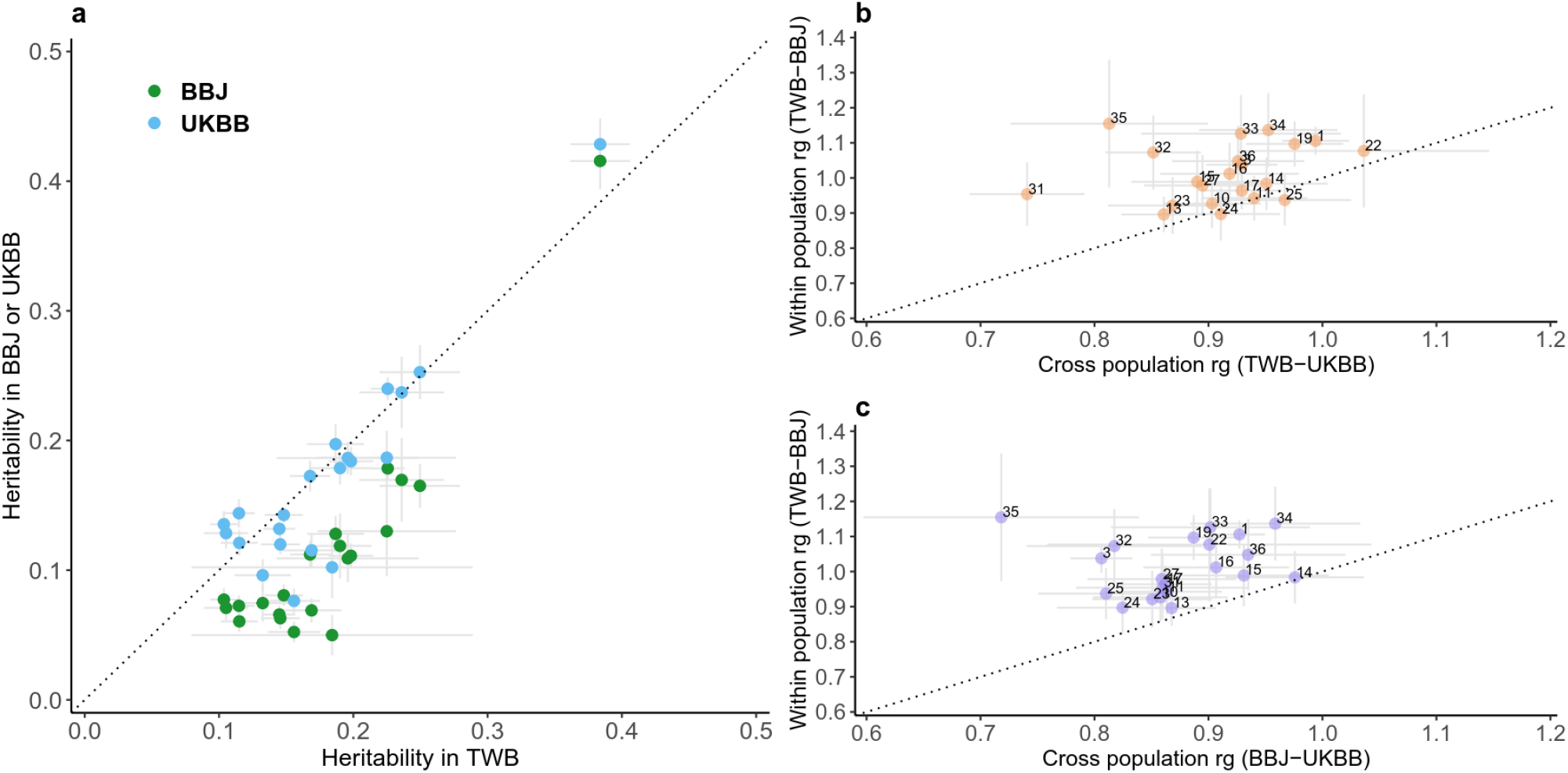
Comparison of SNP-based heritability and within- and cross-population genetic correlation estimates for 21 quantitative traits in TWB, BBJ and UKBB. **a** Comparison of the SNP-based heritability estimates (h^2^_g_) in BBJ or UKBB against TWB. **b** Comparison of the genetic correlation estimates (r_g_) between TWB and BBJ (within EAS) against the cross-population r_g_ estimates between TWB and UKBB (EAS vs. EUR). **c** Comparison of the genetic correlation estimates (r_g_) between TWB and BBJ (within EAS) against the cross-population r_g_ estimates between BBJ and UKBB (EAS vs. EUR). 21 traits for which GWAS summary statistics were available across the three biobanks were included for comparison: HT (1), BMI (3), DBP (10), SBP (11), WBC (13), RBC (14), HB (15), HCT (16), PLT (17), CR (19), T-BIL (22), ALT (23), AST (24), GGT (25), ALB (27), FG (31), HBA1C (32), TC (33), HDL-C (34), LDL-C (35), and TG (36) (See Figure 1 for the abbreviations of full names). All h^2^_g_ and r_g_ were estimated using LD score regression based on high-quality variants available across the three biobanks and GWAS generated by linear regression. The dotted line indicates the diagonal line in each plot. The complete results of the h^2^_g_ and r_g_ analyses are available in Supplementary Tables 7 & 8.

To maximize the power for genetic discovery in EAS populations, we meta-analyzed the GWAS from TWB and BBJ for 23 traits, using an inverse-variance-weighted fixed-effect approach^14^. We report a signal in the meta-analysis as novel if none of the variants within the locus reached genome-wide significance (*P*-value < 5×10^−8^) in BBJ and UKBB GWAS. We identified a total of 2,491 loci associated with the 23 traits, among which 579 were novel (Figure 4a; Supplementary Table 9). For the 13 traits for which BBJ GWAS were not available, we identified an additional 484 genome-wide significant loci using TWB samples only, among which 400 were novel (i.e., no variant in these genetic loci reached genome-wide significance in UKBB) (Supplementary Table 9). The minor allele frequencies (MAF) of the lead SNPs in these 979 novel loci were significantly greater in EAS relative to EUR (average MAF = 30% in EAS vs. 22% in EUR; paired *t*-test *P*-value < 2×10^−16^). As expected, many of the associated loci were highly pleiotropic (Figure 4b; Supplementary Table 10). For example, *TRPS1* was associated with 14 traits spanning the anthropometric, bone, hematological and metabolic categories. The mechanism underlying these pleiotropic associations (e.g., biological pleiotropy vs. mediated pleiotropy) warrants further investigations.

**Figure 4:**
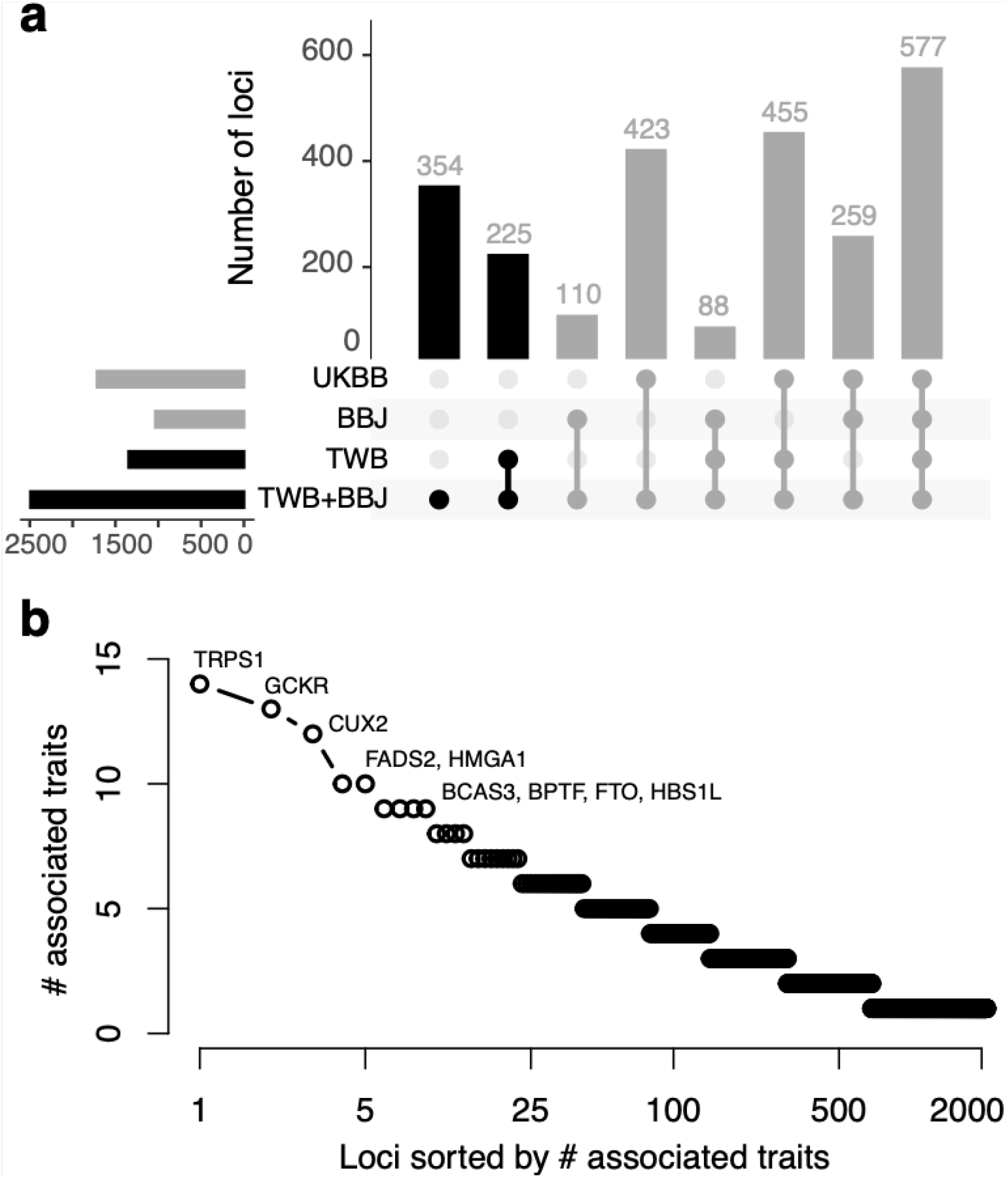
Genetic loci associated with quantitative traits in the East Asian populations. **a** Genome-wide significant loci identified in the TWB and BBJ meta-analysis, tallied based on their significance in TWB, BBJ and UKBB. **b** Distribution of pleiotropic loci defined as the number of associated traits for each locus. **c** The complete results of loci discovery and information on the pleiotropic genes are available in Supplementary Tables 9 & 10.

To assess the clinical utility of biomarker GWAS, we examined whether polygenic risk scores (PRS) of biomarkers can be used to predict the risk of common complex disease. We applied PRS-CSx^24,25^, a Bayesian polygenic prediction method, to integrate the GWAS summary statistics of EAS and EUR ancestry, and calculate both an EAS-specific and an EUR-specific PRS for each biomarker. We then predicted five complex diseases [obesity (Ncase = 824; defined as BMI >= 30), overweight (Ncase = 3,873; defined as BMI >= 25), hypertension (Ncase = 1,149), hyperlipidemia (Ncase = 771), and type 2 diabetes (Ncase = 508)], in a held-out sample of the TWB (N = 10,285; TWBv2 array), using a linear combination of PRS from one or more biomarkers (Figure 5), controlling for age, sex and top 20 principal components (PCs) of genotype data. Biomarker PRS were significantly associated with disease status, explaining >8% of the variation for obesity, overweight and hypertension, 6% of the variation for type 2 diabetes, and 4.3% of the variation for hyperlipidemia on the liability scale (AUC = 0.62 - 0.67; Figure 5; Supplementary Table 11). The odds ratios comparing individuals in the top 10% vs. the remaining 90% of the PRS distribution ranged from 2.0 to 2.7. As a comparison, we also predicted type 2 diabetes using the largest-to-date disease GWAS in EAS (Ncase = 41,223; Ncontrol = 243,023) and EUR (Ncase = 74,124; Ncontrol = 824,006)^26^. While the disease GWAS represented much larger sample sizes, prediction accuracy achieved by biomarker PRS was comparable (Figure 5; Supplementary Table 11). The relative contribution of each biomarker to the prediction of disease risk is summarized in Supplementary Table 12.

**Figure 5:**
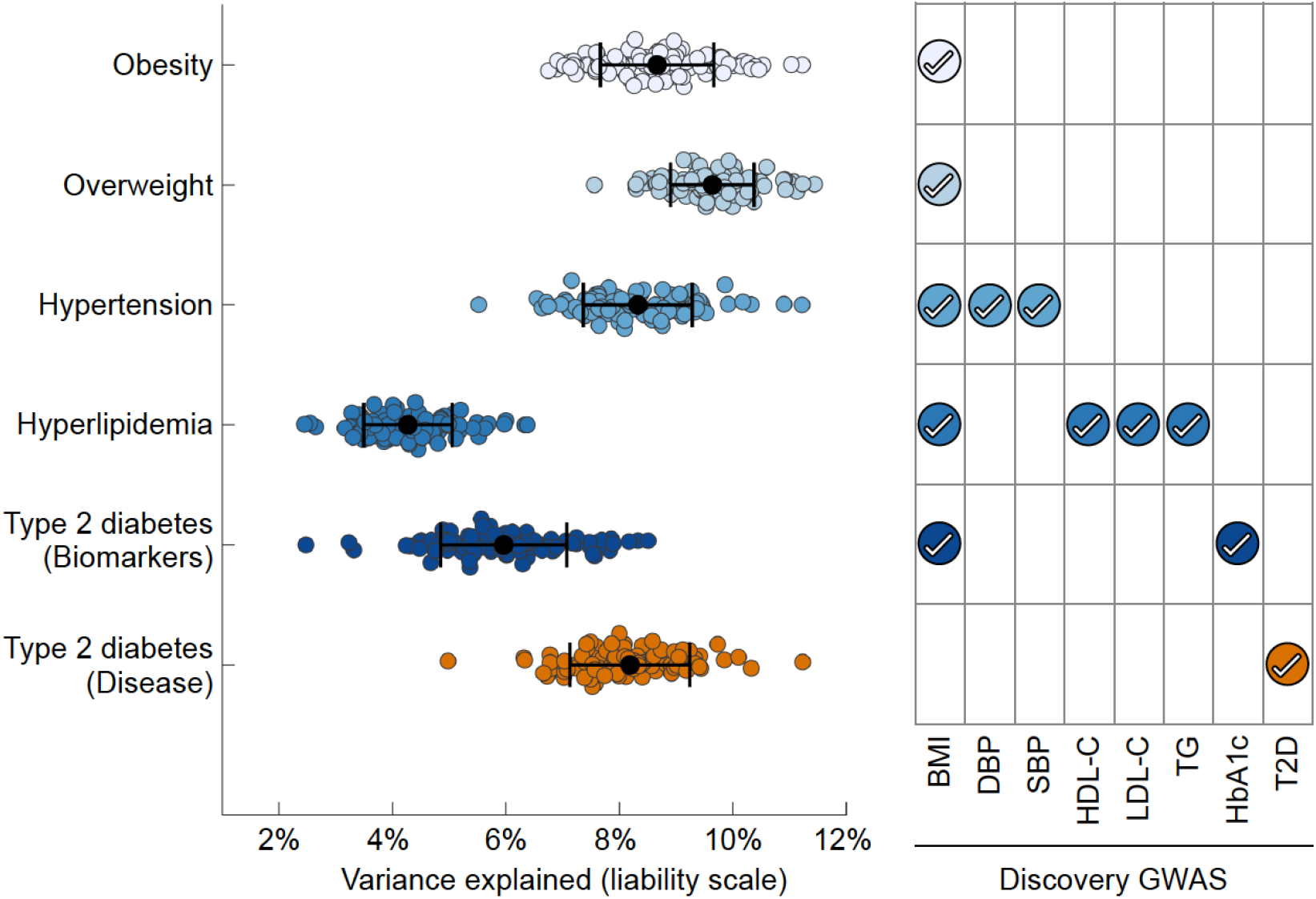
Polygenic prediction of common complex diseases in the Taiwan Biobank. PRS-CSx was applied to jointly model the East Asian (EAS) and European (EUR) GWAS summary statistics of each biomarker, and derive an EAS-specific and an EUR-specific polygenic risk score (PRS). Each disease was predicted by the linear combination of PRS from one or more biomarkers (right panel), controlling for age, sex and top 20 principal components (PCs) of genotype data. The left-out TWB sample (N=10,285) was repeatedly and randomly divided into a validation dataset (where tuning parameters and the optimal linear combination of PRS were learnt), and a testing dataset (where the predictive performance of the final PRS was assessed). To benchmark the predictive performance of biomarker PRS, self-reported type 2 diabetes (T2D) was also predicted by PRS derived from the EAS and EUR type 2 diabetes GWAS. Biomarker GWAS in EAS were obtained from the meta-analysis of TWB and BBJ (N = 115,405 to 230,899); biomarker GWAS in EUR were obtained from UKBB (N = 315,133 to 344,182) and the GIANT study (for BMI; N = 681,275). The T2D disease GWAS in EAS has 41,223 cases and 243,023 controls (4,609 cases and 87,873 controls from TWB; 36,614 cases and 155,150 controls from BBJ); the T2D GWAS in EUR has 74,124 cases and 824,006 controls. Each dot in the left panel represents the prediction accuracy (variance explained on the liability scale) from one random split of the dataset. Error bar represents the standard error of the prediction accuracy across 100 random splits for each disease. PRS performance metrics and the contribution of each PRS to the prediction of disease risk are available in Supplementary Tables 11 & 12.

In summary, we performed genome-wide analysis on 102,900 community-based TWB participants across 36 human complex traits. Leveraging GWAS summary statistics from BBJ and UKBB, we found that the genetic architecture for the quantitative traits examined was largely consistent within EAS and between EAS and EUR populations. Integrating TWB and BBJ GWAS identified a total of 2,975 genetic loci, among which 979 had not been reported in previous biobank GWAS. We additionally fine-mapped over 200 association signals to a single variant with PIP > 95%, and identified 24 putative causal missense variants that implicated novel genes underlying quantitative traits. Lastly, our PRS analysis demonstrated the potential utility of biomarker GWAS in predicting disease risk and the promise of multi-trait cross-population polygenic prediction. We note that, despite the large number of novel loci identified, the current study only included quantitative traits, due to the ongoing data collection efforts on disease phenotypes in TWB, and is thus limited in the scope of phenotypes analyzed. In addition, the collective sample size of TWB and BBJ remains relatively small compared with genetic studies in EUR populations, and may be underpowered for gene discovery of polygenic traits. We also note that we were only able to perform fine-mapping on the TWB sample, rather than using the TWB-BBJ meta-GWAS, due to the limited genomic coverage of the BBJ GWAS; the BBJ sample was imputed to 1KG phase 1 data, which only covers 45% of the variants in the phase 3 reference panel (37.9 million vs. 84.4 million)^5,16^. Nevertheless, the novel findings from genome-wide association analyses, fine-mapping, and polygenic disease risk prediction based on the current analysis in TWB represent a major advance in diversifying GWAS samples and the characterization of the genetic architecture of human complex traits in EAS populations. Future endeavors on increasing the sample size and phenotype coverage in TWB, and improving cross-biobank data harmonization will further facilitate genomic discovery.

## Data Availability

Publicly available data were downloaded from the following databases: 1000 Genomes Project phase 3 data: https://mathgen.stats.ox.ac.uk/impute/1000GP_Phase3.html; BBJ summary statistics: http://jenger.riken.jp/en/result; UKBB summary statistics: http://www.nealelab.is/uk-biobank (GWAS round 2 was used in this study).

## Author contributions

Conceptualized the study: CYC, YAF, HH, TG, YFL

Performed the analysis: TTC, SCL, YFL

Critically reviewed the analysis plan and provided analytic pipelines and scripts: CYC, YAF, RL, HH, TG

Wrote the paper: CYC, YAF, HH, TG, YFL

Critically revised the paper: SHW, YHH, HIY, PHK, MJD, WJC

All authors reviewed and approved the final version of the manuscript.

## Competing interests

Chia-Yen Chen is an employee of Biogen. Yen-Chen Anne Feng is an employee of Vertex Pharmaceuticals. Mark J. Daly is a founder of Maze Therapeutics. The remaining authors declare no competing interests.

## Acknowledgements

We thank Masahiro Kanai for helpful discussions related to Biobank Japan (BBJ). We thank the Neale Lab and BBJ for releasing the genome-wide association summary statistics. Y.F.L. is supported by the National Health Research Institutes (NP-109-PP-09), and the Ministry of Science and Technology (109-2314-B-400-017) of Taiwan. T.G. is supported by NIA K99/R00AG054573. H.H. acknowledges support from NIDDK K01DK114379, NIMH U01MH109539, Brain & Behavior Research Foundation Young Investigator Grant, the Zhengxu and Ying He Foundation, and the Stanley Center for Psychiatric Research. This research has been conducted using the Taiwan Biobank resource. We thank the National Center for Genome Medicine of Taiwan for the technical support in genotyping. We thank the National Core Facility for Biopharmaceuticals (NCFB, MOST 106-2319-B-492-002) and National Center for High-performance Computing (NCHC) of National Applied Research Laboratories (NARLabs) of Taiwan for providing computational and storage resources.

## Ethics

The access to and use of the Taiwan Biobank data in the present work was approved by the Ethics and Governance Council (EGC) of Taiwan Biobank (approval number: TWBR10907-05) and the Institutional Review Board (IRB) of National Health Research Institutes, Taiwan (approval number: EC1090402-E). The data collection of Taiwan Biobank was approved by the Ethics and Governance Council (EGC) of Taiwan Biobank and the Department of Health and Welfare, Taiwan (Wei-Shu-I-Tzu NO.1010267471). Taiwan Biobank obtained informed consent from all participants for research use of collected data. Only publicly available GWAS summary statistics, without individual-level information, were used from Biobank Japan (BBJ) and UK Biobank (UKBB).

## Supplementary Tables

**Supplementary Table 1**: Information on the 36 quantitative traits examined.

**Supplementary Table 2**: Summary of genome-wide significant loci for the Fuji plot.

**Supplementary Table 3**: Heritability estimates within discovery batch 1 and 2, and genetic correlation estimates between the two discovery batches of the Taiwan Biobank (TWB) samples across 36 quantitative traits.

**Supplementary Table 4**: Heritability estimates across 36 quantitative traits in the Taiwan Biobank (TWB).

**Supplementary Table 5**: Pairwise genetic correlation estimates between 36 quantitative traits in the Taiwan Biobank (TWB).

**Supplementary Table 6**: Full results for the fine-mapping analysis.

**Supplementary Table 7**: Heritability estimates across 21 quantitative traits within the Taiwan Biobank (TWB), Biobank Japan (BBJ) and UK Biobank (UKBB).

**Supplementary Table 8**: Cross-biobank genetic correlation estimates across 21 quantitative traits.

**Supplementary Table 9**: Information on the novel loci identified by combining data from the Taiwan Biobank (TWB) and Biobank Japan (BBJ).

**Supplementary Table 10**: Information on the pleiotropic genes.

**Supplementary Table 11**: Performance metrics for the polygenic prediction of five common complex diseases.

**Supplementary Table 12**: Relative contribution of each polygenic risk score (PRS) to the prediction of five common complex diseases.

## METHODS

### Taiwan biobank (TWB)

#### Sample characteristics, quality control and Imputation

The Taiwan Biobank (TWB) (https://www.twbiobank.org.tw/new_web_en/) is a prospective cohort study of the Taiwanese population with multi-omics genomic data and repeated measurements of a wide range of phenotypes collected from 149,894 individuals (as of April 2021), with an expected final sample size of 200,000. Participants in TWB were recruited across 29 recruitment centers with at least one center in each city or county of Taiwan. TWB has two recruiting arms: the community-based arm and the hospital-based arm. All participants included in this study were from the community-based arm. TWB collects extensive phenotypes including demographics, socioeconomic status, environmental exposures, lifestyle, dietary habits, family history and self-reported disease status through structured questionnaires. The anthropometric measures, and blood and urine samples were collected at recruitment, and biomarkers were assayed subsequently, following the manufacturer’s protocol at Linkou, Taiwan. TWB participants were 30 to 70 years old at recruitment, and the sex ratio was 0.57 in the final analytic sample of this study (37,225 males and 65,675 females; see Figure 1).

We obtained genome-wide genotype data from a total of 110,926 TWB participants. Genotyping was performed using two different customized chips. 27,719 samples were genotyped on the TWBv1 custom array, which was designed based on Thermo Fisher Axiom Genome-Wide CHB Array with customized contents; 83,207 samples were genotyped on the TWBv2 custom array, which was designed by Thermo Fisher Scientific Inc. based on whole-genome sequencing data from 946 TWB samples with customized contents.^10^ We divided the samples genotyped on the TWBv2 array into two subsets, with 68,975 samples for loci discovery and 14,232 samples for polygenic risk score (PRS) analysis. We refer to samples genotyped on the TWBv1 array as “discovery batch 1” and the loci discovery samples genotyped on the TWBv2 array as “discovery batch 2” (see Figure 1).

We conducted stringent quality control (QC) for the two discovery samples and the sample for PRS analysis separately before imputation^4,27^. QC was performed using a combination of bash script, R, python and PLINK^4,27^, with scripts adapted from a recent biobank genotype QC project: https://github.com/Annefeng/PBK-QC-pipeline. We first filtered out variants with call rate < 0.98 and samples with call rate < 0.98, and removed variants that were duplicated, monogenic or not correctly mapped to a genomic position. We then merged TWB samples with the 1000 Genomes (1KG) Project phase 3 data (N=2,504)^16^, and selected high-quality, common variants by removing multi-allelic and strand ambiguous SNPs, SNPs with call rate < 0.98 and minor allele frequency (MAF) < 5%, and SNPs located in long-range LD regions (chr6: 25-35Mb; chr8: 7-13Mb). Next, we performed LD-pruning at *R*^2^ = 0.1, and computed principal components (PCs) of the merged genotype data with LD-pruned variants. Using the population labels of 1KG samples as the reference, we trained a random forest model with top 6 PCs to classify TWB samples into 1KG super-population groups: East Asian [EAS], European [EUR], African [AFR], American [AMR], South Asian [SAS]. We retained TWB samples that can be assigned to a homogeneous EAS population group with a predicted probability of EAS ancestry > 0.8. After initial population assignment, we filtered out samples with heterozygosity rate outside of 6 standard deviation (SD) from the sample average, and samples with mismatched genetic and self-reported sex. We then performed three rounds of in-sample principal components analysis (PCA) to identify remaining population outliers, each time removing TWB samples with any of the top 10 PCs that was more than 6 SD away from the sample average. We used the in-sample PCs derived after outlier removal in subsequent analyses. Lastly, we removed variants with call rate < 0.98 and Hardy-Weinberg equilibrium (HWE) test *P*-value < 1e-10 within the EAS sample.

After pre-imputation QC, we used Eagle v2.4^28^ for pre-phasing and Minimac4^29^ for genotype imputation with 1KG phase 3 data as the reference panel. We performed post-imputation QC by retaining variants with imputation INFO > 0.6 and MAF > 0.5% for the downstream analyses, which included up to 8,190,806 variants for discovery batch 1 and 8,156,315 variants for discovery batch 2, respectively. The imputed dataset included a total of 92,615 samples for loci discovery (27,033 samples for discovery batch 1 and 65,582 samples for discovery batch 2), and 12,997 samples for polygenic risk score (PRS) analysis. We further restricted the PRS analysis to 10,285 unrelated individuals that were also unrelated with the discovery GWAS samples. We note that while most of the phenotypes examined in this study were measured on the large majority of the discovery samples, the final analytic sample size of GWAS reduced to 62,901 (17,111 from discovery batch 1 and 45,790 from discovery batch 2) for FEV1, FVC and FEV1R, and 65,360 (19,509 from discovery batch 1 and 45,851 from discovery batch 2) for AFP due to missing phenotypic data.

#### Genetic association analysis

We performed genetic association analysis on 36 quantitative traits including anthropometric measures and biomarkers from 8 categories as described in Figure 1. The three repeated measures for systolic and diastolic blood pressure and resting heart rate within the same visit were averaged. For each trait, we removed samples with phenotypic measures that were more than 6 SD away from the sample average. By doing so, we also removed samples with extremely high alpha-fetoprotein (AFP) levels that could be a result of pregnancy. We also note that the bone mineral density was measured using ultrasound at heel and then converted to T-score (BMD-T) using an Asian young adult reference and Z-score (BMD-Z) using an Asian age-matched reference. In addition, we randomly removed one sample from each pair of duplicated samples within and across the two discovery batches. We then performed inverse rank-based normal transformation (IRNT) within each batch to achieve normality of the phenotype. All following genetic analyses were based on IRNT transformed measures. Association analyses were performed separately for the two discovery batches, followed by an inverse-variance-weighted fixed-effect meta-analysis (based on effect size estimates and standard errors) implemented in METAL^14^.

We used linear regression implemented in Regenie^11^ for association testing, controlling for age, age^2^, sex, age by sex interaction, age^2^ by sex interaction, and top 20 PCs. Regenie is a two-step whole-genome regression approach that accounts for potential population stratification and sample relatedness, providing better statistical power than restricting the association analysis to unrelated individuals. Specifically, in step 1 of Regenie, we used a subset of LD-pruned (at *R*^2^ = 0.9) SNPs with imputation INFO > 0.8 and MAF > 1% (919,630 SNPs for discovery batch 1 and 745,794 SNPs for discovery batch 2) to calculate a leave-one-chromosome-out (LOCO) polygenic score for each trait and each individual using Ridge regression. Association testing was then performed, in step 2 of Regenie, including the LOCO polygenic predicted value from step 1 as an offset in the linear regression model, in addition to other covariates, to account for sample relatedness. Association tests were performed in three sets of phenotypes according to missing data patterns: (1) AFP; (2) FEV1, FVC, FEV1R; (3) all other phenotypes. The association test statistics were then meta-analyzed between discovery batch 1 and 2. The final meta-analysis results only included variants presented on both batches.

### Biobank Japan (BBJ)

All Biobank Japan (BBJ) GWAS summary statistics were publicly available: http://jenger.riken.jp/en/result^5–7^. We included 23 GWAS of quantitative traits from BBJ: height (HT), body mass index (BMI), diastolic blood pressure (DBP), systolic blood pressure (SBP), white blood cell (WBC), red blood cell (RBC), hemoglobin (HB), hematocrit (HCT), platelet (PLT), blood urea nitrogen (BUN), creatinine (CR), uric acid (UA), total bilirubin (T-BIL), alanine aminotransferease (ALT), aspartate aminotransferase (AST), gamma-glutamyltransferase (GGT), albumin (ALB), fasting glucose (FG), hemoglobin A1c (HbA1c), total cholesterol (TC), high-density-lipoprotein cholesterol (HDL-C), low-density-lipoprotein cholesterol (LDL-C), and triglyceride (TG). Phenotypes used in these GWAS were either converted to z-scores (HT, TG, DBP, SBP, WBC, RBC, HB, HCT, PLT, BUN, CR, UA, AST, ALT, GGT, TC, HDL-C, LDL-C) or inverse rank-based normal transformated (BMI, T-BIL, ALB, FG, HbA1c). All association tests were performed by first residualizing phenotypes on age, age^2^, sex, top 10 PCs, and trait-specific covariates (e.g., disease status), followed by a linear regression, except for height for which association tests were conducted with a linear mixed-effects model implemented in BOLT-LMM (v2.2).^6,30^ We meta-analyzed each of these 23 GWAS with the corresponding GWAS in TWB using sample-size-weighted z-score meta-analysis implemented in METAL.^14^ We retained variants presented in either TWB or BBJ in this meta-analysis for loci discovery in the East Asian populations (Figure 4; Supplementary Tables 9 & 10).

### UK Biobank (UKBB)

All UK Biobank GWAS summary statistics used in this study were publicly available: http://www.nealelab.is/uk-biobank. These GWAS were conducted and released by Benjamin Neale’s lab at Massachusetts General Hospital and the Broad Institute. We included 27 GWAS from UKBB: height (HT), weight (WT), body mass index (BMI), body fat rate (BFR), waist circumference (WC), diastolic blood pressure (DBP), systolic blood pressure (SBP), white blood cell (WBC), red blood cell (RBC), hemoglobin (HB), hematocrit (HCT), platelet (PLT), creatinine (CR), Microalbumin urine (mALB), total bilirubin (T-BIL), alanine aminotransferease (ALT), aspartate aminotransferase (AST), gamma-glutamyltransferase (GGT), albumin (ALB), forced expiratory flow (FEV1), forced vital capacity (FVC), fasting glucose (FG), hemoglobin A1c (HbA1c), total cholesterol (TC), high-density-lipoprotein cholesterol (HDL-C), low-density-lipoprotein cholesterol (LDL-C), and triglyceride (TG). All phenotypes used in these GWAS were inverse rank-based normal transformated. Association tests were conducted using linear regrerssion, controlling for age, age^2^, sex, age by sex interaction, age^2^ by sex interaction, and top 20 PCs.

### Pleiotropic genes

To identify genes influencing multiple traits, we took the list of genes mapped to each locus from FUMA^15^ (“genes.txt” from the FUMA download), and picked the gene with the most significant *P*-value in the “minGwasP” column for each locus. Multiple genes were retained for a locus if they shared the minimal *P*-value. We then report the number of traits a gene was associated with in the “Number of trait” column in Supplementary Table 10, and visualized the distribution of pleiotropic loci in Figure 4b.

### Fine-mapping

We implemented a summary statistics based version of SuSiE (Sum of Single Effects)^17^ in Python for the fine-mapping analysis (Code availability). All loci identified through FUMA using the TWB summary statistics were extended +/-100kb to ensure sufficient locus data was available for fine-mapping. In-sample LD was calculated using hard-called genotypes merged across post-imputation samples from the two discovery batches. We identified the 95% credible set using the following settings: marginal *P*-value threshold < 5×10^−8^, minimum purity = 0.5, algorithmic convergence tolerance = 10^−4^, and the maximum number of iterations = 100. An initial run was performed limiting to a maximum of 5 signals. Loci in which five credible sets were identified were re-run, relaxing the maximum number of signals to 10. Loci that failed to converge in the initial run were rerun through an iterative process of reducing the maximum number of signals from 5 towards 1 until model convergence. Annotations for credible sets were generated using ANNOVAR^31,32^ (version 2019Oct24) on the GENCODE V19 database, and ExAC using VEP version 101^31,32^.

### Heritability and genetic correlation analyses

LD score regression (LDSC)^13^ was derived under linear regression and thus may produce biased heritability (h^2^_g_) and genetic correlation (r_g_) estimates when applied to GWAS summary statistics generated from mixed models. In addition, all association tests in BBJ and UKBB were performed under linear regression (except for height in BBJ). Therefore, to enable a fair comparison of h^2^_g_ and r_g_ between TWB, BBJ and UKBB, we applied LDSC to GWAS summary statistics generated from linear regression in unrelated TWB samples; the sample size for h^2^_g_ and r_g_ analysis (ranged from 53,962 to 79,407; Supplementary Tables 3, 4 & 5) was thus smaller than the Regenie-based GWAS sample size. Specifically, we removed one sample in each pair of second degree or more closely related relatives within and across the two discovery batches in TWB, and performed association tests in the remaining unrelated individuals, controlling for age, age^2^, sex, age by sex interaction, age^2^ by sex interaction, and top 20 PCs, followed by meta-analysis across the two batches using PLINK2.^27^ This set of association GWAS results was used in within-TWB h^2^_g_ and r_g_ estimation shown in Figure 2b & 2c and Supplementary Tables 3, 4 & 5, and cross-biobank h^2^_g_ and r_g_ comparisons presented in Figure 3 and Supplementary Tables 7 & 8.

We used LDSC to estimate h^2^_g_ for 36 traits and the pairwise r_g_ between them within TWB. We additionally applied LDSC to BBJ and UKBB GWAS summary statistics to estimate h^2^_g_ for 21 traits that were available across the three biobanks. For fair comparison, all LDSC analyses were restricted to shared SNPs with INFO > 0.8 and MAF > 1% across the three biobanks. LD scores were calculated using the 1KG phase 3 reference panel that matched the ancestry of the GWAS sample. We note that the h^2^_g_ estimate for total bilirubin (T-BIL) had an unusually large standard error (SE = 0.105; median SE for the other traits = 0.016; see Figure 2 and Supplementary Table 7). We identified a Mendelian locus for T-BIL on chromosome 2 (*P*-value < 1e-1000), harboring the *UGT1A1* gene, which is known to cause inherited unconjugated hyperbilirubinemia, including Gilbert Syndrome (OMIM: 143500), Crigler–Najjar Syndrome type I (OMIM: 218800), and Crigler–Najjar Syndrome type II (OMIM: 606785). Removing this Mendelian locus reduced LDSC h^2^_g_ estimate from 0.184 to 0.091 and its SE from 0.105 to 0.020. We separately estimated the T-BIL phenotypic variance explained by the top signal in the Mendelian locus as a fixed effect to be 0.088, which matched the missing h^2^_g_ by removing this locus from the LDSC analysis.

We used the baseline-LD-X model of S-LDXR^22^ (version 0.3-beta) to estimate cross-biobank genetic correlations, using shared SNPs with INFO > 0.8 and MAF > 1% across the three biobanks. To estimate the cross-population r_g_ between TWB and UKBB, and between BBJ and UKBB, we used the default LD scores for EAS and EUR populations provided by S-LDXR as the reference panels. To estimate within-EAS r_g_ between TWB and BBJ, we used the LD scores and regression weight files for EAS provided by S-LDXR as the reference panels for both biobanks.

### Polygenic prediction

For each biomarker (BMI, DBP, SBP, HDL-C, LDL-C, TG, and HbA1c), we collected the largest GWAS in EAS (the meta-analysis of TWB and BBJ) and EUR populations (Neale Lab UKBB GWAS for all biomarkers except BMI, for which GWAS summary statistics from the GIANT study^33^ were used). Population-specific PRS for each biomarker was calculated using PRS-CSx^25^, a Bayesian polygenic prediction method that jointly models GWAS summary statistics from multiple populations to improve polygenic prediction. Specifically, for a fixed global shrinkage parameter (phi = 1e-6, 1e-4, 1e-2, and 1.0 in this study) that models the overall sparseness of the genetic architecture, PRS-CSx returned posterior SNP effect size estimates for each discovery population (i.e., EAS and EUR), which were used to calculate both an EAS-specific PRS and an EUR-specific PRS in the left-out TWB sample (N=10,285) that was unrelated to the discovery samples in TWB. We predicted five common complex diseases (obesity, defined as BMI>=30; overweight, defined as BMI>=25; hypertension; hyperlipidemia, and type 2 diabetes) using the PRS of one or more biomarkers. Specifically, we predicted obesity and overweight using BMI, hypertension using BMI, DBP and SBP, hyperlipidemia using BMI, HDL-C, LDL-C and TG, and type 2 diabetes using BMI and HbA1c (Figure 5, right panel). We repeatedly and randomly divided the left-out TWB sample into a validation dataset (N=5,000), where we selected the optimal global shrinkage parameter for each biomarker and learnt the optimal linear combination of the PRS across biomarkers that were used as predictors, and a testing dataset (N=5,285), where we evaluated the predictive performance of the final PRS, controlling for age, sex and top 20 PCs. This process was repeated for 100 times. To compare the prediction accuracy of PRS derived from biomarker GWAS and disease GWAS, we additionally applied PRS-CSx to the largest EAS (the meta-analysis of TWB and BBJ) and EUR type 2 diabetes GWAS^26^, and used the resulting PRS to predict self-reported type 2 diabetes in the same TWB held-out sample.

## Data availability

Publicly available data were downloaded from the following databases: 1000 Genomes Project phase 3 data: https://mathgen.stats.ox.ac.uk/impute/1000GP_Phase3.html; BBJ summary statistics: http://jenger.riken.jp/en/result; UKBB summary statistics: http://www.nealelab.is/uk-biobank (“GWAS round 2” was used in this study).

## Code availability

Regenie: https://github.com/rgcgithub/regenie;

PLINK2: https://www.cog-genomics.org/plink/2.0;

METAL: https://genome.sph.umich.edu/wiki/METAL;

FUMA: https://fuma.ctglab.nl;

Fuji plot: https://github.com/mkanai/fujiplot;

LDSC: https://github.com/bulik/ldsc;

S-LDXR: https://huwenboshi.github.io/s-ldxr;

PRS-CSx: https://github.com/getian107/PRScsx;

Python implementation for SuSiE (attached as a tarball and will be released in GitHub upon publication)

## Notes

### Author Declarations

Taiwan Biobank obtained ethical approval from the Ethics and Governance Council (EGC) of Taiwan Biobank and the Department of Health and Welfare, Taiwan (Wei-Shu-I-Tzu NO.1010267471). Taiwan Biobank obtained informed consent from all participants. The Taiwan Biobank approved an application for access to and use of the data (TWBR10907-05), and ethical approval for the analysis was obtained from the Institutional Review Board (IRB) of National Health Research Institutes, Taiwan (EC1090402-E).

## References

1. Visscher, P. M. et al. 10 Years of GWAS Discovery: Biology, Function, and Translation. Am. J. Hum. Genet. 101, 5–22 (2017).

2. Tam, V. et al. Benefits and limitations of genome-wide association studies. Nat. Rev. Genet. 20, 467–484 (2019).

3. Martin, A. R. et al. Clinical use of current polygenic risk scores may exacerbate health disparities. Nat. Genet. 51, 584–591 (2019).

4. Peterson, R. E. et al. Genome-wide Association Studies in Ancestrally Diverse Populations: Opportunities, Methods, Pitfalls, and Recommendations. Cell 179, 589–603 (2019).

5. Kanai, M. et al. Genetic analysis of quantitative traits in the Japanese population links cell types to complex human diseases. Nat. Genet. 50, 390–400 (2018).

6. Akiyama, M. et al. Characterizing rare and low-frequency height-associated variants in the Japanese population. Nat. Commun. 10, 4393 (2019).

7. Akiyama, M. et al. Genome-wide association study identifies 112 new loci for body mass index in the Japanese population. Nat. Genet. 49, 1458–1467 (2017).

8. Sudlow, C. et al. UK biobank: an open access resource for identifying the causes of a wide range of complex diseases of middle and old age. PLoS Med. 12, e1001779 (2015).

9. Bycroft, C. et al. The UK Biobank resource with deep phenotyping and genomic data. Nature 562, 203–209 (2018).

10. Wei, C.-Y. et al. Genetic profiles of 103,106 individuals in the Taiwan Biobank provide insights into the health and history of Han Chinese. NPJ Genom Med 6, 10 (2021).

11. Mbatchou, J. et al. Computationally efficient whole genome regression for quantitative and binary traits. bioRxiv (2020) doi:10.1101/2020.06.19.162354.

12. Bulik-Sullivan, B. K. et al. LD Score regression distinguishes confounding from polygenicity in genome-wide association studies. Nat. Genet. 47, 291–295 (2015).

13. Bulik-Sullivan, B. et al. An atlas of genetic correlations across human diseases and traits. Nat. Genet. 47, 1236–1241 (2015).

14. Willer, C. J., Li, Y. & Abecasis, G. R. METAL: fast and efficient meta-analysis of genomewide association scans. Bioinformatics 26, 2190–2191 (2010).

15. Watanabe, K., Taskesen, E., van Bochoven, A. & Posthuma, D. Functional mapping and annotation of genetic associations with FUMA. Nat. Commun. 8, 1826 (2017).

16. 1000 Genomes Project Consortium et al. A global reference for human genetic variation. Nature 526, 68–74 (2015).

17. Wang, G., Sarkar, A., Carbonetto, P. & Stephens, M. A simple new approach to variable selection in regression, with application to genetic fine mapping. J. R. Stat. Soc. Series B Stat. Methodol. 82, 1273–1300 (2020).

18. Polfus, L. M. et al. Whole-Exome Sequencing Identifies Loci Associated with Blood Cell Traits and Reveals a Role for Alternative GFI1B Splice Variants in Human Hematopoiesis. Am. J. Hum. Genet. 99, 785 (2016).

19. Johnson, A. D. et al. Genome-wide association meta-analysis for total serum bilirubin levels. Hum. Mol. Genet. 18, 2700–2710 (2009).

20. Chambers, J. C. et al. Genome-wide association study identifies loci influencing concentrations of liver enzymes in plasma. Nat. Genet. 43, 1131–1138 (2011).

21. Shi, H. et al. Localizing Components of Shared Transethnic Genetic Architecture of Complex Traits from GWAS Summary Data. Am. J. Hum. Genet. 106, 805–817 (2020).

22. Shi, H. et al. Population-specific causal disease effect sizes in functionally important regions impacted by selection. Nat. Commun. 12, 1098 (2021).

23. Lam, M. et al. Comparative genetic architectures of schizophrenia in East Asian and European populations. Nat. Genet. 51, 1670–1678 (2019).

24. Ge, T., Chen, C.-Y., Ni, Y., Feng, Y.-C. A. & Smoller, J. W. Polygenic prediction via Bayesian regression and continuous shrinkage priors. Nat. Commun. 10, 1776 (2019).

25. Ruan, Y. et al. Improving polygenic prediction in ancestrally diverse populations. medRxiv (2021) doi:10.1101/2020.12.27.20248738.

26. Mahajan, A. et al. Fine-mapping type 2 diabetes loci to single-variant resolution using high-density imputation and islet-specific epigenome maps. Nat. Genet. 50, 1505–1513 (2018).

27. Chang, C. C. et al. Second-generation PLINK: rising to the challenge of larger and richer datasets. Gigascience 4, 7 (2015).

28. Loh, P.-R. et al. Reference-based phasing using the Haplotype Reference Consortium panel. Nat. Genet. 48, 1443–1448 (2016).

29. Das, S. et al. Next-generation genotype imputation service and methods. Nat. Genet. 48, 1284–1287 (2016).

30. Loh, P.-R. et al. Efficient Bayesian mixed-model analysis increases association power in large cohorts. Nat. Genet. 47, 284–290 (2015).

31. Wang, K., Li, M. & Hakonarson, H. ANNOVAR: functional annotation of genetic variants from high-throughput sequencing data. Nucleic Acids Res. 38, e164 (2010).

32. Lek, M. et al. Analysis of protein-coding genetic variation in 60,706 humans. Nature 536, 285–291 (2016).

33. Yengo, L. et al. Meta-analysis of genome-wide association studies for height and body mass index in ∼700000 individuals of European ancestry. Hum. Mol. Genet. 27, 3641–3649 (2018).

